# Iron related biomarkers predict disease severity in a cohort of Portuguese adult patients during COVID-19 acute infection

**DOI:** 10.1101/2021.09.09.21263251

**Authors:** Ana C. Moreira, Maria J. Teles, Tânia Silva, Clara M. Bento, Inês Simões Alves, Luísa Pereira, João T. Guimarães, Graça Porto, Pedro Oliveira, Maria Salomé Gomes

## Abstract

**BACKGROUND:** Growing evidence indicates a link between iron metabolism and COVID-19 clinical progression, supporting the use of iron and inflammatory parameters as relevant biomarkers to predict patients’ outcomes.

**METHODS:** We evaluated iron metabolism and immune response in 303 patients admitted to the main hospital of the northern region of Portugal with variable clinical pictures, from September to November 2020. Of these, 127 tested positive for SARS-CoV-2 and 176 tested negative. Iron-related laboratory parameters and cytokines were determined in blood samples collected soon after admission and, in a subgroup of patients, throughout hospitalization. Demographic data, comorbidities and clinical outcomes were recorded. Patients were assigned into 5 groups according to disease severity.

**RESULTS:** Serum iron and transferrin levels at admission were lower in COVID-19-positive than in COVID-19-negative patients. Conversely, the levels of interleukin(IL)-6 and monocyte chemoattractant protein 1 (MCP1) were increased in COVID-19-positive patients. The lowest serum iron and transferrin levels at diagnosis were associated with the worst outcomes. Iron levels negatively correlated with IL-6 and higher levels of this cytokine were associated with a worse prognosis. Serum ferritin levels at diagnosis were higher in COVID-19-positive than in COVID-19-negative patients but did not correlate with disease severity. Longitudinal determinations of iron and ferritin made in a subgroup of patients (n=23) revealed highly variable results.

**CONCLUSIONS:** Serum iron is the simplest laboratory test to be implemented as a predictor of disease progression in hospitalized acute COVID-19-positive patients. Variation of ferritin with time should be revisited in larger cohorts.

**Key points:** COVID-19-positive patients have lower serum iron and higher ferritin than COVID-19-negative patients in variable clinical contexts. Lowest serum iron and highest IL-6 levels at hospital admission associate with the poorest outcomes. Association of serum ferritin with disease progression is debatable.

## INTRODUCTION

The clinical spectrum of COVID-19 is wide, ranging from slight fatigue to severe acute respiratory failure and death [1,2]. The identification of relevant early biomarkers that can predict disease progression and severity is very important to allow a more efficient allocation of resources, to identify and protect groups at higher risk, and to reduce mortality.

Alterations of iron distribution are important host defense mechanisms against infection [3-5]. The most consistently reported alteration of iron distribution in COVID-19-positive patients is hypoferremia [6-8]. Hypoferremia in the context of infection and inflammation is usually attributed to hepcidin [9]. However, contradictory results have been obtained regarding the correlations between iron or hepcidin levels and disease severity [10-14]. Another iron-related alteration that has been proposed as predictor of disease severity in COVID-19 is hyperferritinemia [6,10,11,13-17]. However, its impact on disease prognosis is not consistent [6-8].

In our study, we compared iron related, inflammatory and hematological biomarkers in COVID-19-positive patients with COVID-19-negative patients admitted to the same hospital during the same period. The results support the notion that infection by SARS-CoV-2 elicits a distinctive pattern of alterations in the host immune response and iron homeostasis and stresses the importance of implementing the use of simple iron related parameters in the analytical workup of these patients.

## METHODS

### Study design

We analyzed data from 127 patients diagnosed with COVID-19 at Centro Hospitalar e Universitário de São João (CHUSJ), Porto, Portugal, between September 2, and November 17, 2020. COVID-19 diagnosis was based on SARS-CoV-2 RNA detection by reverse-transcription real-time PCR in a nasopharyngeal swab sample. Blood samples collected in the context of clinical evaluation were used to obtain routine analytical data and to make additional determinations relevant to this study, as explained below. The interval between the SARS-CoV-2 PCR test and the first blood collection was less than 6 days for all patients except for 4 positive patients. These came to the hospital several days after a diagnosis was made elsewhere, making the interval between diagnosis and the first sample increase to 8, 10, 11 and 21 days, respectively.

For comparison, samples from 176 patients admitted to the hospital with variable clinical pictures in the same period, with a negative test for SARS-CoV-2 RNA, were also investigated. All patients 18 years old or more were considered eligible, except those with active or recent oncological disease, pregnancy, or hematological disease. Additionally, 35 serum samples were obtained from blood donors at the Clinic Hematology Service of Centro Hospitalar Universitário do Porto (CHUP) in the same metropolitan area as CHUSJ. Blood donors were not tested for SARS-CoV-2, but they had no symptoms of disease and filled the criteria for blood donation, thus were used as healthy controls.

The study was approved by the local ethics committee of CHUSJ and performed in the respect of the Helsinki declaration. Routine clinical data were collected in an anonymized format and no additional procedures or sample collections were performed for this study. As such, the study was considered by the ethical committee as exempt from the need to take specific written informed consent from the patients enrolled.

### Patient stratification

Once enrolled, patients were followed according to Hospital standards. According to the severity of the disease, patients were assigned to one of 5 groups: 1) asymptomatic, 2) outpatients 3) general inward, 4) intensive care unit, 5) fatal outcome. The criteria for hospital stay or ambulatory, as well as for admission at a general ward or an intensive care unit, were based on clinical signs and symptons and were the same all over the study period. Each patient was assigned to the group corresponding to the worst condition experienced during the follow-up for that specific clinical condition.

### Laboratory determinations

Complete blood counts and erythrocytes parameters were determined using a Sysmex XE-2100 or XE-5000 (Emilio de Azevedo Campos, Portugal) equipment, in fresh peripheral blood anticoagulated with K3-EDTA. Serum or heparin-plasma samples were used to determine erythropoietin, iron, transferrin, ferritin, and C-reactive protein. Erythropoietin was determined using a chemiluminescent immunoassay on an Immulite 2000 analyzer (Siemens Healthcare Diagnostics). Iron and transferrin were determined by a colorimetric method and ferritin by an immunoturbidimetric assay, using Olympus reagents and analyzer (Olympus Diagnostics). Normal reference ranges were obtained from National Health Authorities’ Regulations [18] and from commercial companies, according to the hospital laboratory procedures.

Heme quantification was performed using the QuantiChrom Heme Assay Kit from BioAssay Systems (Hayward, California, USA). Hepcidin levels were measured using Hepcidin 25 (bioactive) HS ELISA from DRG instruments GmbH (Marburg, Germany). Haptoglobin was measured using Human Haptoglobin ELISA kit from Abcam, plc (Cambridge, United Kingdom). Quantification of cytokines was performed with the BD cytometric bead array (CBA) human inflammation kit (catalog no. 551811) from BD Biosciences (San Jose, CA, USA), or with Human Inflammation Panel 1 Mix and Match Subpanel Catalog. No. 740809 from BioLegend (San Diego, CA, USA). Data were analyzed with BD Accuri C6 with FCAP Array software (BD Biosciences) or Legendplex software (San Diego, CA, USA), respectively. The lower detection limits were: IL-1beta: 0.5pg/ml, IFN alpha 2: 0.8pg/ml, IFN gamma: 1.0pg/ml), TNF alpha: 0.2pg/ml, IL-6: 0.5pg/ml, IL-8 3.0pg/ml, IL-10: 0.9pg/ml, IL-12p70: 0.2pg/ml, IL-18: 1.0pg/ml, IL-23: 1.0pg/ml, IL-33: 10pg/ml, MCP-1: 5pg/ml.

### Statistical analyses

For quantitative variables with pronounced asymmetry, data are presented by median, P25 and P75. Otherwise, data are presented as mean ± standard deviation. Categorical variables are presented by their counts and corresponding percentages. When comparing two groups, an unpaired t-test or Main Whitney test was performed. For comparisons between groups, one-way-ANOVA was performed after confirming the normality and homogeneity of variances and the Kruskal Wallis test was used, when these assumptions were not met. Post hoc analysis was performed by Tukey HSD test. Correlation coefficients were obtained using Spearman’s rho. Association between variables was assessed by Chi-square test or Fisher’s exact test. Statistical significance was reached when p value was lower than 0.05 unless otherwise specified. Statistical analysis was performed using SPSS from IBM, USA.

## RESULTS

### Baseline characterization of the patients

As detailed in Methods, we conducted an observational study of patients admitted to CHUSJ most of them with symptoms compatible with COVID-19. According to the result of the PCR test for SARS-CoV-2, the patient was included in the -positive or -negative group. Blood was collected typically within the first 48h after admission. Sera from healthy blood donors were used for comparisons. Table 1 summarizes the baseline demographic and clinical characterization of the individuals enrolled. The number and type of co-morbidities was similar between COVID-19-positive and COVID-19-negative patients. The most prevalent co-morbidity was hypertension, followed by dyslipidemia. Diabetes was also present in an important proportion of the patients, especially among COVID-19-positive. This picture reflects the overall prevalence of these morbidities in the Portuguese population [19].

**Table 1.** Baseline demographic and clinical characterization of the individuals included in the study.

|  | COVID19-negative |  | COVID19-positive |
| --- | --- | --- | --- |
|  | Blood donors | Patients |  |
| Number of patients | 35 | 176 | 127 |
| Median Age (P<sub>25</sub>-P<sub>75</sub>)<sup>a</sup> | 46 (42-59) | 65 (50-78) | 72 (61-81) |
| Number of Males (%) | 16 (45.7) | 102 (58.0) | 78 (61.4) |
| Number of comorbidities, Median (P<sub>25</sub>-P<sub>75</sub>)<sup>a</sup> |  | 4.0 (2-6) | 4.0 (2-6) |
| Frequency of comorbidities (%): |  |  |  |
| Hypertension (*) |  | 52.8 | 65.4 |
| Dyslipidemia (ns) |  | 44.3 | 44.1 |
| Diabetes (**) |  | 26.1 | 40.9 |
| Obesity (ns) |  | 23.3 | 24.4 |
| Chronic Kidney Disease (ns) |  | 15.3 | 14.2 |
| Chronic Respiratory Disease (ns) |  | 13.6 | 14.2 |
| Iron Supplementation (ns) |  | 10.8 | 5.5 |
| Hypocoagulation (ns) |  | 9.7 | 8.7 |
| Anemia (ns) |  | 3.4 | 3.1 |

<sup>a</sup>Median (percentile 25-percentile 75) for age and number of comorbidities; \*p<0.05; \*\*p<0.01; ns: not significant

### Impact of COVID-19 in iron-related and hematological parameters

Both COVID-19-positive and COVID-19-negative patients had lower serum iron at Hospital admission than healthy blood donors (Figure 1a). Moreover, COVID-19-positive patients had significantly lower serum iron levels than COVID-19-negative patients (25 (17-42) versus 42 (23.3-76.8) µg/dL). Defining hypoferremia as serum iron level lower than 50 µg/dL, 80.7% of COVID-19-positive and 57.1% (p<0.001) of COVID-19-negative patients were hypoferremic at admission, suggesting that COVID-19 and serum iron levels are not independent. Accordingly, transferrin level was also lower in COVID-19-positive compared to COVID-19-negative patients (Figure 1b). Of note, decreased transferrin in COVID-19-positive patients was not related to hepatic disorder, as 91.5% of the patients with low transferrin levels had alanine aminotransferase concentration below twice the highest normal value. Conversely, the levels of serum ferritin were significantly higher in COVID-19-positive (628.2 (365.8-1246.0) ng/mL) as compared to COVID-19-negative patients (190.5 (119.9-382.4) ng/mL) and blood donors (56.9 (34.3-127.3) ng/mL) (Figure 1d). Interestingly, hepcidin levels were not significantly different between COVID-19-positive (73.3 (32.8-103.9) nM) and COVID-19-negative patients (36.52 (6.0-118.3) nM). However, in both cases they were significantly higher than in blood donors (14.5 (2.4-32) nM) (Figure 1e). No significant differences were found between blood donors, COVID-19-positive and COVID-19-negative patients, regarding heme, haptoglobin or erythropoietin levels (Figure 1f,g,h).

**Figure 1.**
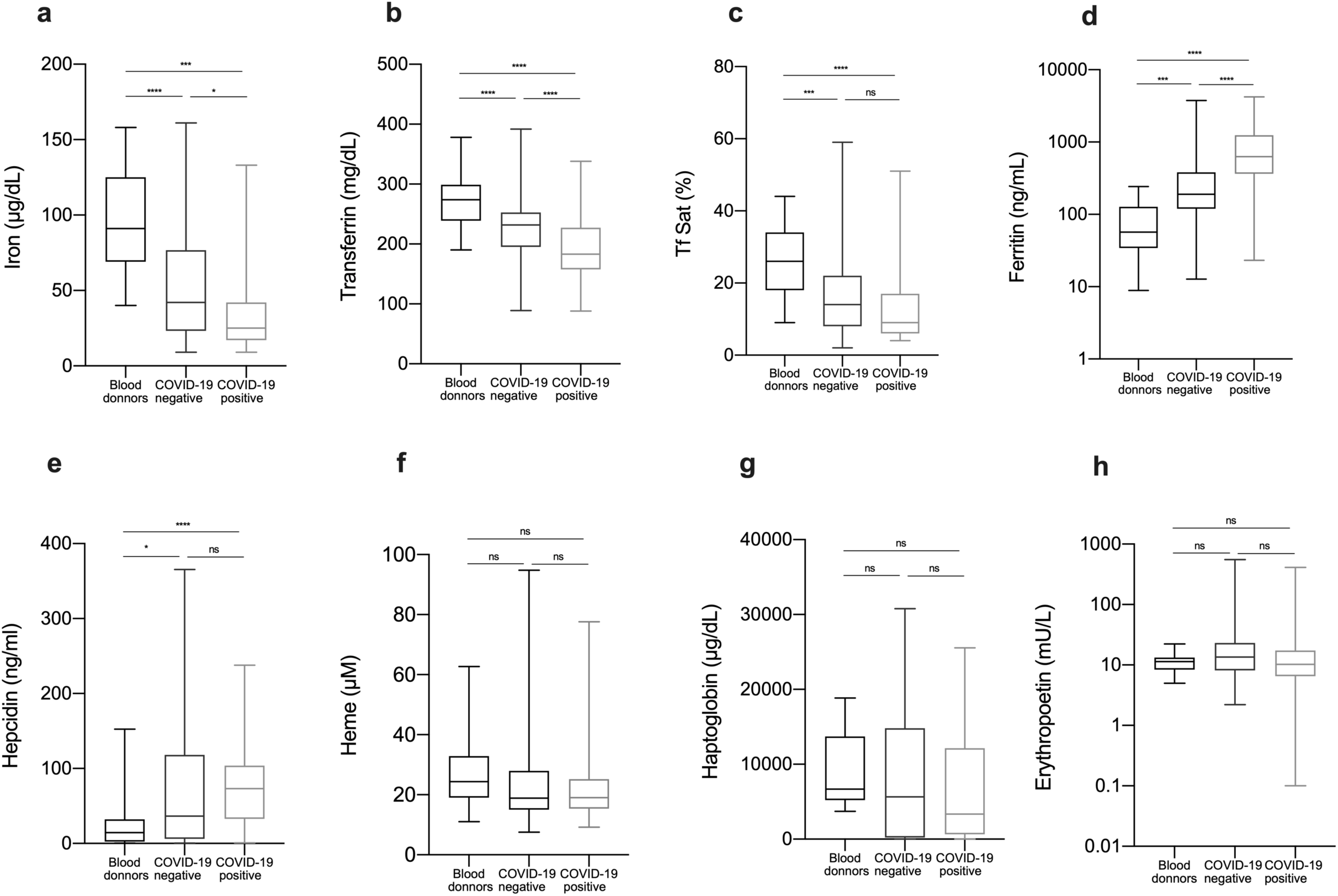
Blood iron parameters are altered in COVID-19 patients. Blood samples were obtained at patients’ admission to CHUSJ and plasma or serum was used to measure iron, transferrin, transferrin saturation, ferritin, erythropoietin, heme, haptoglobin, and hepcidin. Samples obtained from healthy blood donors were used for comparison. Statistical analysis was performed with one-way-ANOVA and Kruskal Wallis test, following multiple comparisons with Sidak and Dunn’s test respectively. *p<0.05, **p<0.01, ***p<0.001

Despite the clear decrease in serum iron levels, COVID-19-positive patients on average had no decrease in hemoglobin concentration or red blood cells (RBC) counts, when compared to COVID-19-negative patients (Table 2).

**Table 2.** Hematological and biochemical determinations on the first sample after hospital admission

|  |  | Normal range | COVID-19-negative | COVID-19-positive |  |
| --- | --- | --- | --- | --- | --- |
| Red blood cells (10<sup>12</sup>/L) |  |  |  |  |  |
|  | Males | 4.31-6.4 | 4.4 ± 0.80 | 4.5 ± 0.63 | ns |
|  | Females | 3.85-5.20 | 4.1 ± 0.60 | 4.3 ± 0.54 | * |
| Hemoglobin (g/dL) |  |  |  |  |  |
|  | Males | 13.6-18 | 13.3 ± 2.62 | 13.6 ± 2.03 | ns |
|  | Females | 11.5-16 | 12.1 ± 1.93 | 12.73 ± 1.61 | ns |
| Hematocrit (%) |  |  |  |  |  |
|  | Males | 39.8-52 | 39.3 ± 7.08 | 40.1 ± 5.67 | ns |
|  | Females | 34.7-46 | 36.44 ± 5.30 | 38.4 ± 4.57 | * |
| Mean corpuscular volume (fL) |  | 80 -97 | 89.7 ± 6.28 | 90.1 ± 5.55 | ns |
| Mean corpuscular hemoglobin (pg) |  | 26 – 34 | 30.0 ± 2.49 | 30.2 ± 2.14 | ns |
| Mean corpuscular hemoglobin concentration (g/dL) |  | 32 – 36 | 33.5 ± 1.43 | 33.6 ± 1.15 | ns |
| Red blood cell distribution width-CV (%) |  | 11.5 – 15 | 14.2 ± 1.68 | 13.8 ± 1.60 | ns |
| Red blood cell distribution width-SD (fL) |  | 37 – 54 | 46.1 ± 5.99 | 45.4 ± 5.38 | ns |
| White blood cells (x10<sup>9</sup>/L) |  | 4.0 – 10.0 | 9.4 ± 4.20 | 7.0 ± 2.94 | *** |
| Neutrophils (x10<sup>9</sup>/L) |  | 1.5 – 8 | 6.9 ± 3.96 | 5.3 ± 2.80 | *** |
| Lymphocytes (x10<sup>9</sup>/L) |  | 0.8 – 4 | 1.6 ± 1.24 | 1.1 ± 0.61 | *** |
| Monocytes (x10<sup>9</sup>/L) |  | 0.0 – 1.2 | 0.7 ± 0.36 | 0.5 ± 0.31 | *** |
| Platelets (10<sup>9</sup>/L) |  | 140 - 440 | 220.0 ± 85.61 | 200.8 ± 80.72 | * |
| Mean platelet volume (fL) |  |  | 10.8 ± 1.01 | 10.9 ± 1.03 | ns |
| Platelet distribution width (fL) |  |  | 12.8 ± 2.42 | 13.1 ± 2.37 | ns |
| CRP<sup>a</sup> (mg/L) |  | <3 | 53.6 ± 94.64 | 86.0 ± 81.15 | ** |
| AST<sup>a</sup> (U/L) |  | 10-37 | 40.3 ± 43.84 | 61.3 ± 136.90 | ns |
| ALT<sup>a</sup> (U/L) |  | 10-37 | 31.2 ± 24.40 | 44.7 ± 75.50 | ns |
| Gamma GT<sup>a</sup> (U/L) |  | 10-49 | 73.4 ± 161.01 | 82.4 ± 124.72 | ns |
| Total Protein (g/L) |  | 64-83 | 68.1 ± 9.95 | 69.1 ± 9.32 | ns |

<sup>a</sup>CRP: C-Reactive-Protein; AST: Aspartate Aminotransferase; ALT: Alanine Aminotransferase; Gamma GT: Gamma-glutamyl transferase. Results are presented as mean ± standard deviation and comparisons were performed using unpaired Mann Whitney t-test. Statistical differences: \*p<0.05, \*\*p<0.01, \*\*\* p<0.001, ns=not significant

Although on average the white blood cells, neutrophils, monocytes, lymphocytes and platelets counts were within the intervals considered as normal, COVID-19-positive patients had significantly lower levels than COVID-19-negative. Of note, 36.1% of COVID-19-positive patients were lymphopenic at diagnosis (number of lymphocytes lower than 0.8×10^9^/L), as compared to 14.2% of COVID-19-negative patients (p<0.001), consistent with previous studies [1,16,20].

In general, all patients had increased levels of hepatic enzymes and C-reactive protein (CRP), the latter being significantly higher in COVID19-positive patients (Table 2).

### Relationship between iron-related parameters and disease severity

To further understand whether alterations in iron metabolism were related to disease severity, patients were assigned to one of 5 severity groups, as described in the Methods section and summarized in Table 3. Group 1 included 35 healthy blood donors, as well as 15 patients admitted to the hospital with no symptoms suggestive of COVID-19: 8 had psychiatric disorders (organic causes were excluded), 2 had light gastrointestinal symptoms without signs of acute surgical abdomen or infection 5 came for minor elective surgeries without systemic inflammation or infection. Group 2 included patients with mild clinical conditions, which did not imply hospitalization. Patients included in group 3 had to stay in hospital for at least 1 day. Most patients stayed 7 days, 25% stayed less than 5 days and only 35% stayed more than 11 days. Patients included in group 4 were patients admitted to the intensive care unit. Group 5 corresponds to fatal cases.

**Table 3.** Stratification of individuals according to the severity of disease

|  |  | Number | Minimum O<sub>2</sub> saturation<sup>a</sup> | Pneumonia (%) |
| --- | --- | --- | --- | --- |
| **All** |  |  |  |  |
|  | COVID-19-Negative | 176 | 93.0 ± 7.3 | 15.3% |
|  | COVID-19-Positive | 127 | 88.3 ± 7.9 | 60.6% |
| **1 Asymptomatic** |  |  |  |  |
|  | COVID-19-Negative | 15 | 96.8 ± 2.1 | --- |
|  | COVID-19-Positive | 0 | --- | --- |
| **2 Ambulatory** |  |  |  |  |
|  | COVID-19-Negative | 40 | 95.1 ± 3.4 | 15.0% |
|  | COVID-19-Positive | 28 | 94.4 ± 2.5 | 14.3% |
| **3 General inward** |  |  |  |  |
|  | COVID-19-Negative | 72 | 92.9 ± 5.6 | 20.8% |
|  | COVID-19-Positive | 48 | 89.1 ± 6.9 | 60.4% |
| **4 Intensive Care** |  |  |  |  |
|  | COVID-19-Negative | 43 | 93.0 ± 7.2 | 9.3% |
|  | COVID-19-Positive | 32 | 86.4 ± 7.0 | 84.4% |
| **5 Fatalities** |  |  |  |  |
|  | COVID-19-Negative | 6 | 77.3 ± 18.4 | 33.3% |
|  | COVID-19-Positive | 19 | 81.2 ± 10.0 | 89.5% |

<sup>a</sup>Minimum O<sub>2</sub> saturation values are presented as mean ± standard deviation

Notably, serum iron levels at first evaluation were significantly lower in fatal cases, as compared to survivors, both in COVID-19-positive and in COVID-19-negative patients (Figure 2a). Also, the transferrin levels were lower in fatal cases than in outpatients or general inward patients. Serum transferrin was significantly higher in outpatients (group 2) than in all the groups of inpatients (groups 3, 4 and 5) (Figure 2b). Interestingly, when we compared patients at the same level of disease severity, we found that COVID-19-positive patients always had significantly lower iron and transferrin levels than COVID-19-negative patients (Figure 2a,b), except for serum iron in fatal cases.

In the opposite direction, the levels of ferritin were higher in COVID-19-positive in comparison with COVID-19-negative patients in all severity groups (Figure 2c). However, the levels of ferritin were not significantly different between severity groups. No statistically significant differences were found in hepcidin levels, either between severity groups or between COVID-19-positive and negative patients (not shown).

**Figure 2.**
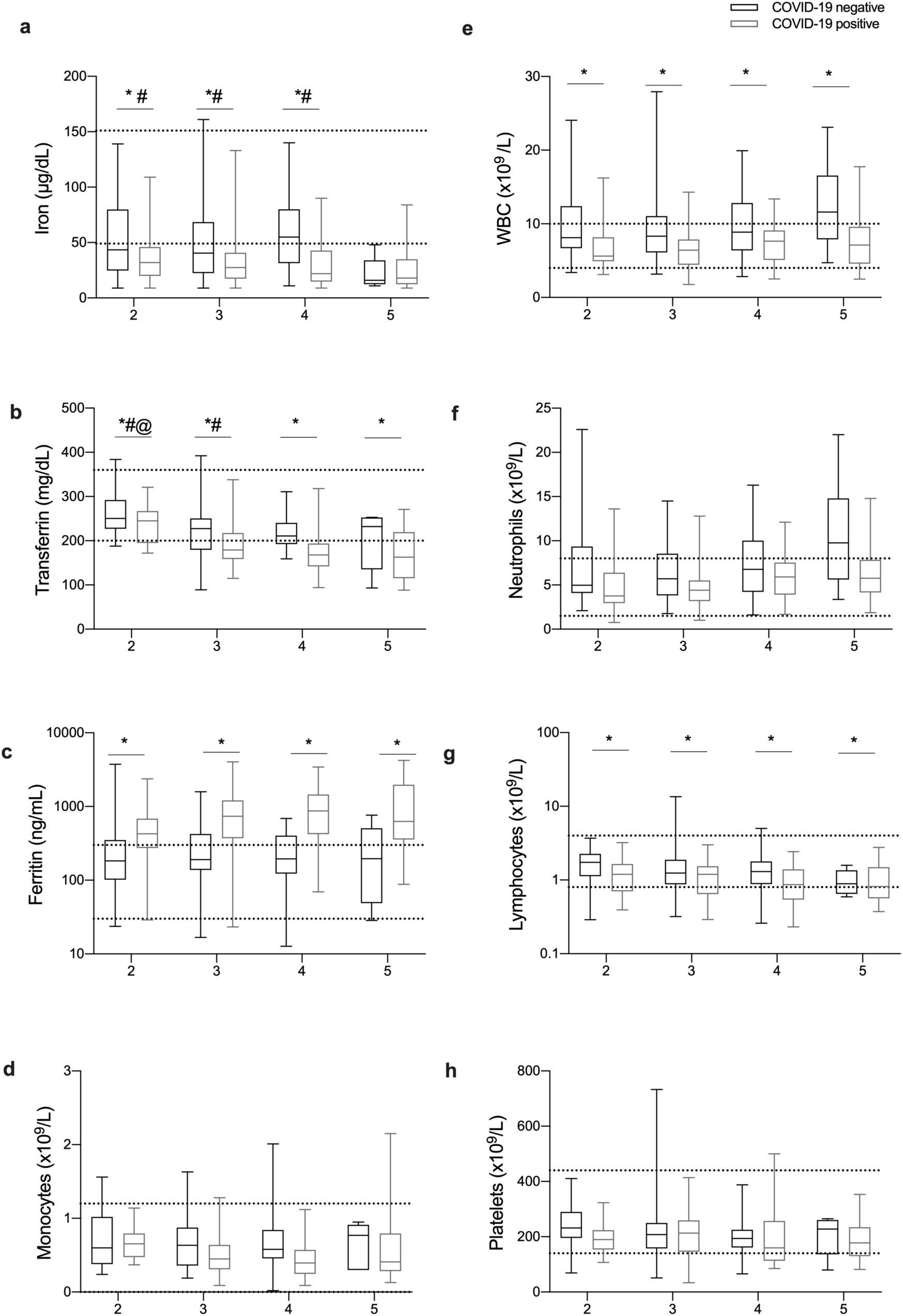
Alterations in blood iron parameters in COVID-19 are related to disease severity. Patients were stratified according to disease severity, as described in the Methods section. The first blood sample available from each patient after admission was used to quantify different types of white blood cells and to measure iron, transferrin and ferritin in plasma or serum. Boxes represent the lower and higher quartile, and the horizontal bar represents the median value. The whiskers represent the minimum and maximum values of each group. Horizontal dotted lines represent maximum and minimum values of the normal range considered at CHUSJ following the National Health Authorities’ Regulations. Statistical analysis was performed with one-way-ANOVA and Kruskal Wallis test, following multiple comparisons with Sidak and Dunn’s test respectively. *p<0.05 between COVID-19-positive and COVID-19-negative patients. #<0.05 compared with severity 5, @<0.05 compared with severities 4 and 5.

Regarding the leukocyte counts, WBC and lymphocytes were lower in COVID-19-positive than in COVID-19-negative patients in all severity levels (Figure 2e,g), while the decreases in monocytes, neutrophils and platelets lost statistical significance after stratification for disease severity (Figure 2d,f,h).

In this cohort, patients with previously diagnosed hypertension, and/or dyslipidemia had an increased chance of developing severe disease (Table S1). Anemia, chronic respiratory disease, chronic kidney disease, diabetes, obesity, or hypocoagulation were not significantly associated with severe forms of the disease. In fact, in this cohort none of the (11) previously hypocoagulated COVID-19-positive patients died (Table S1).

### Inflammation and immune response markers

Iron metabolism is strongly regulated by immune mediators. We measured the serum levels of interferon gamma, interferon alpha 2, TNF alpha, MCP-1, IL-1beta, IL-6, IL-8, IL-10, IL-12p70, IL-18, IL-23 and IL-33. Some of these cytokines were hardly detectable in circulation, although in general they were more frequently detected in samples from COVID-19-positive than in those from COVID-19-negative patients, particularly IL-6 and MCP-1 previously associated to iron metabolism (Table S2).

In Figure 3, we show the quantifications of those cytokines that could be detected in more than 60% of the samples. The levels of circulating IL-6 and MCP-1 were significantly elevated in COVID-19-positive patients when compared to COVID-19-negative patients (Figure 3a,f). Importantly, outpatients (group 2) had lower IL-6 levels than fatal cases (group 5) (Figure 3a). Interestingly, IL-10 was higher on average in COVID-19-positive than in COVID-19-negative patients (32.7 (21.4-61.5) versus 12.7(5.9-28.5)). However, this difference was not observed once the patients were stratified according to severity groups (Figure 3c).

**Figure 3.**
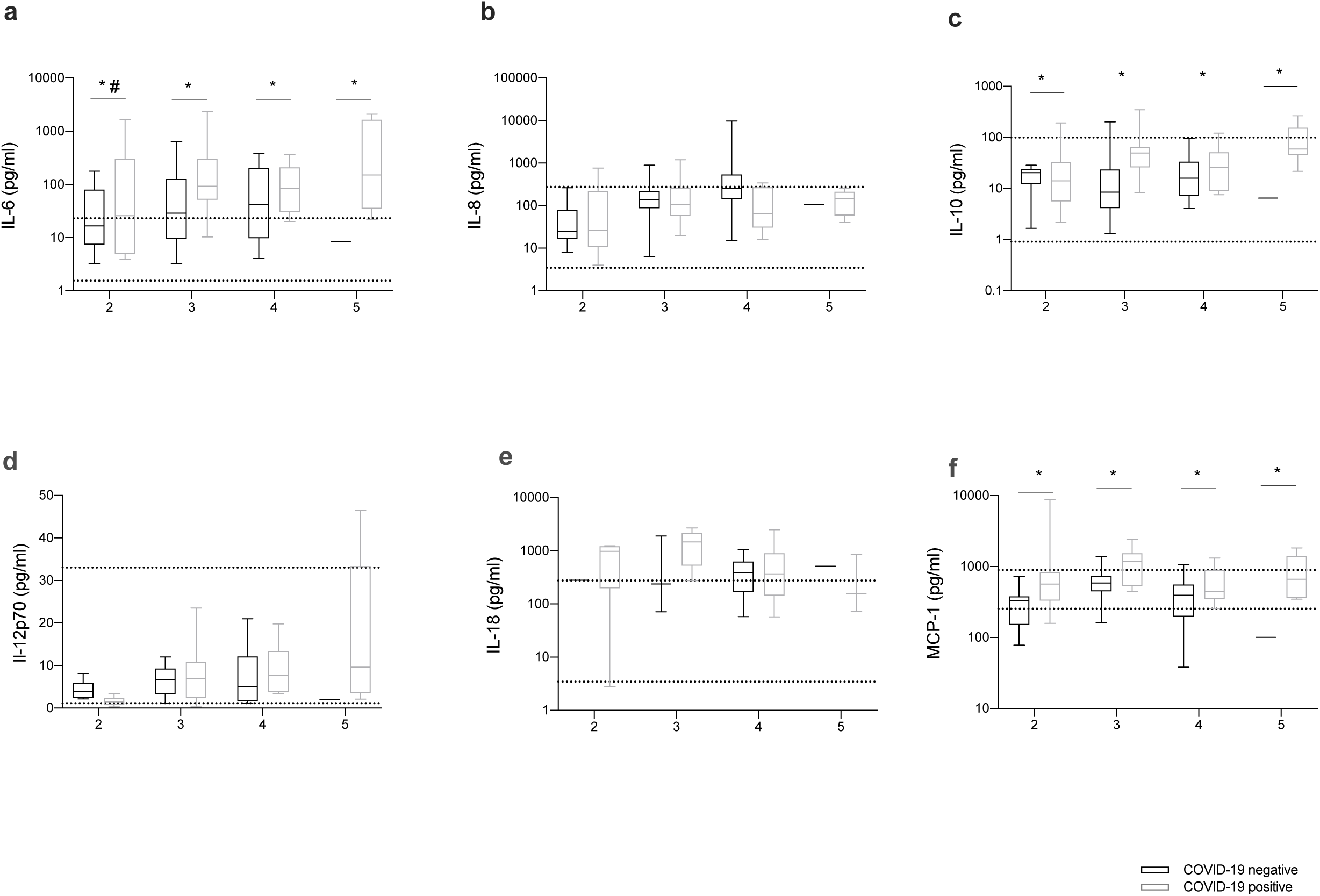
Cytokine levels in COVID-19 are related to disease severity. Patients were stratified according to disease severity, as described in the Methods section. The first blood sample available from each patient after admission was used to quantify IL-6, IL-8, IL-10, IL-12p70, and MCP-1 using cytometric beads assay as described in the methods section. Multiple comparisons were made after one-way-ANOVA. Boxes represent the lower and higher quartile, and the horizontal bar represents the median value. The whiskers represent the minimum and maximum values of each group. Horizontal dotted lines represent the lowest and higher levels measured in blood donors’ samples. Statistical analysis was performed with one-way-ANOVA and Kruskal Wallis test, following multiple comparisons with Sidak and Dunn’s test respectively. *p<0.05 between COVID-19-positive and COVID-19-negative patients, #<0.05 compared with severity 5 within COVID-19-positive patients.

### Correlations between iron parameters and cytokine levels

To gain some insight into the mechanisms of iron dysregulation in COVID-19-positive patients, we performed association tests between specific parameters. We found a significant, although modest, negative correlation between iron and CRP and IL-6, but not other cytokines or hepcidin (Figure 4). Interestingly, the level of transferrin was also negatively correlated with CRP, but also with IL-8, IL-18, hepcidin and ferritin. Ferritin levels were positively correlated with CRP, IL-18 and hepcidin. As expected, a positive correlation was found between hepcidin and IL-6, but also (more surprising) between hepcidin and IL-10. The levels of inflammatory cytokines were positively correlated with each other (Figure 4).

**Figure 4.**
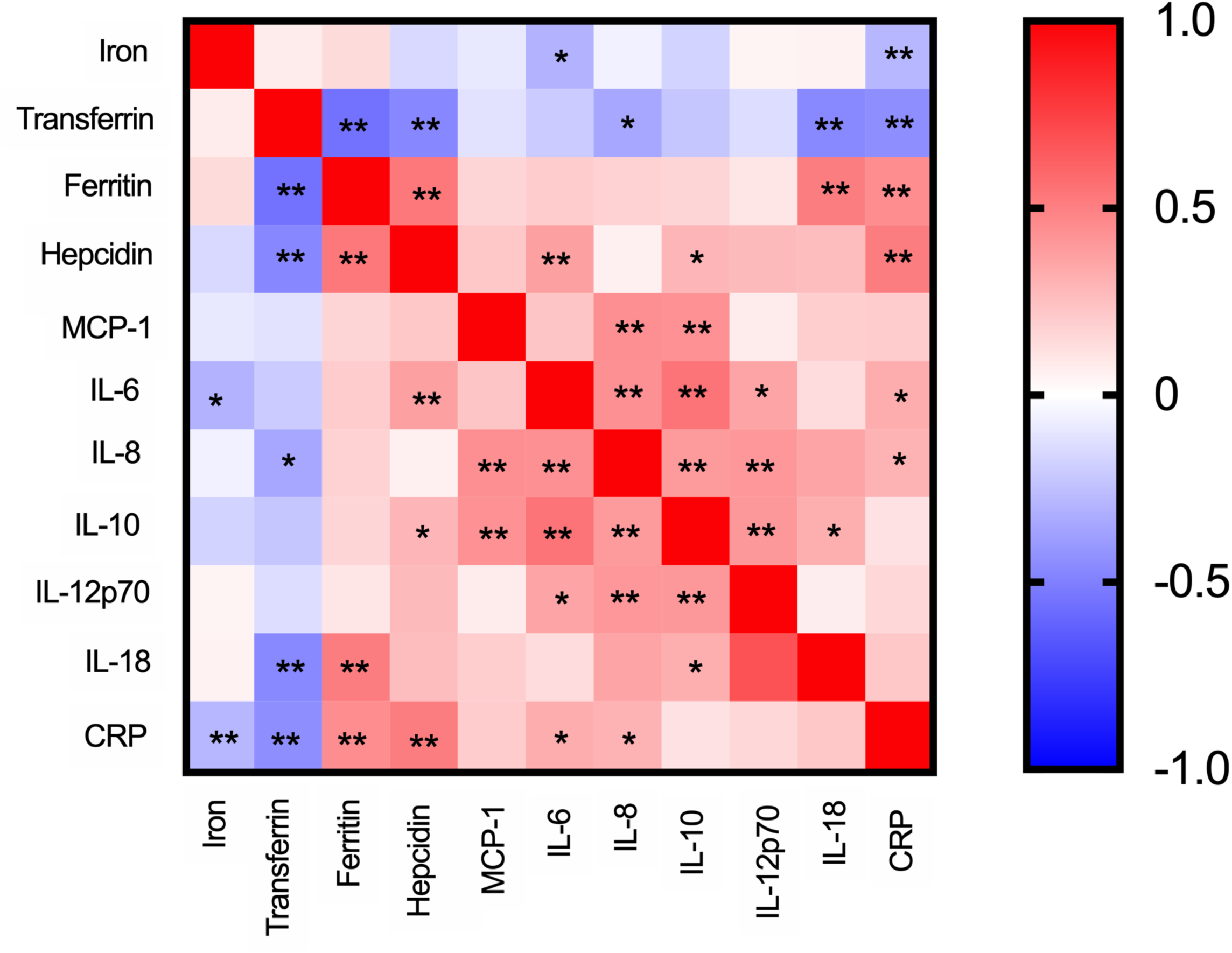
Spearman correlations between different iron markers and cytokines. Non-parametric Spearman correlations were obtained in iron parameters or cytokines determinations of the first sample of the COVID-19-positive patients enrolled in this study. Spearman’s rho was converted in color from blue to red and significance within positive or negative correlations were shown as *p<0.05 and **p<0.01

### Evolution of iron and immune parameters over time

For a limited number of admitted patients, we had access to multiple blood samples along the hospitalization, enabling us to evaluate the evolution of iron and immune-related parameters with time. To achieve some homogeneity, we selected those patients for which the first sample was collected up to 6 days after diagnosis and with at least one sample 4 to 9 days after the first one. Given the low number of patients included (5 fatal cases and 18 survivors) and the high heterogeneity of the observations, no major conclusions could be drawn for most parameters (Figure 5 and Figure S1). However, it was possible to observe a tendency for a decrease in serum iron over time in fatal cases, contrasting with stabilization or increase in recovery cases (Figure 5a). In contrast, serum ferritin tended to increase over time in fatal cases and decrease or stabilize in recovery cases (Figure 5c). Additional studies are needed to draw conclusions.

**Figure 5.**
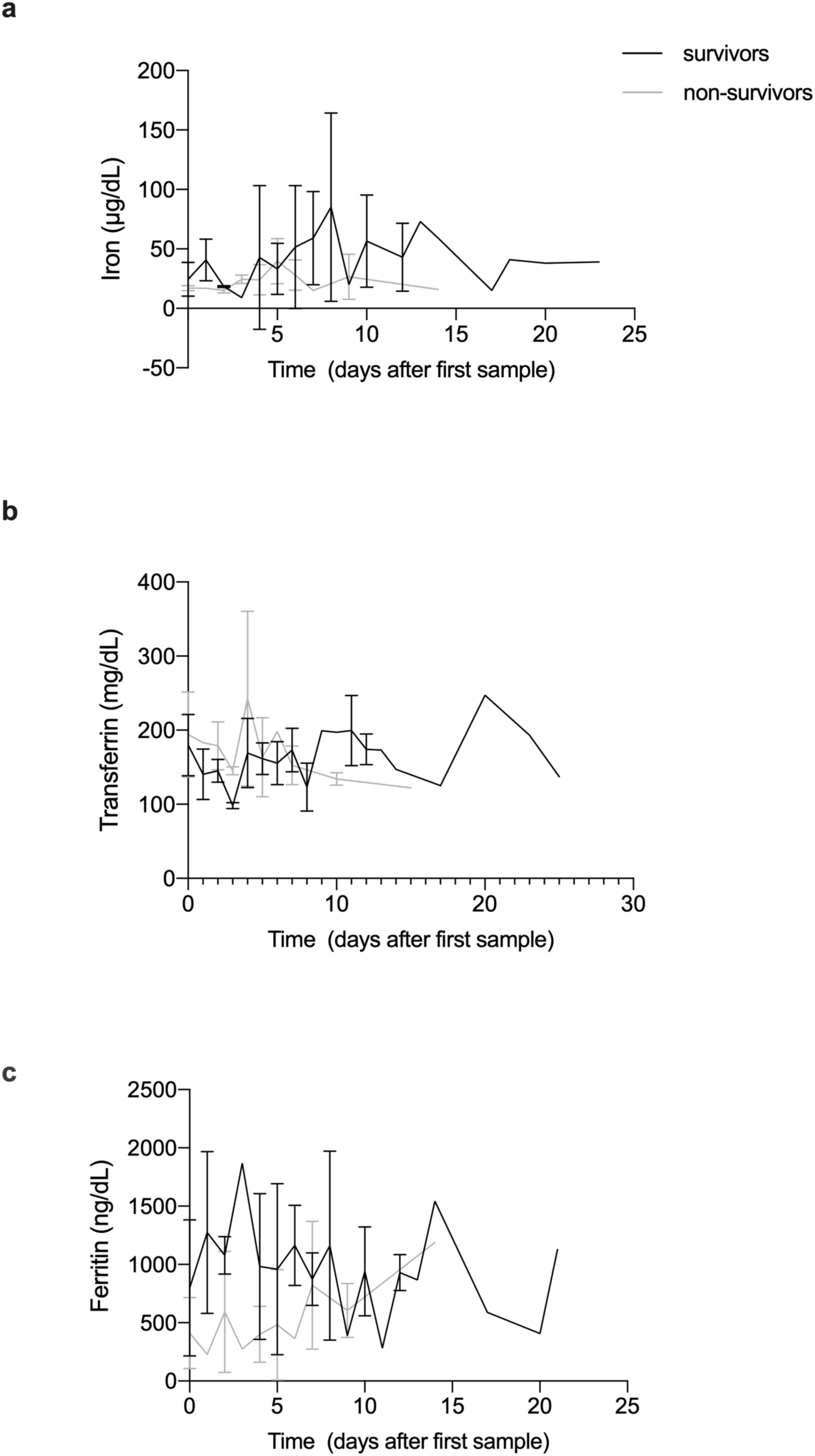
Evolution of iron parameters over time, during hospital stay. Blood samples were obtained over the time of stay, from a limited number of COVID-19-positive inpatients: 5 fatal cases and 18 survivors. Circulating iron levels, transferrin and ferritin were measured as described in the methods section. Graphs show mean and standard deviations within each group: survivors (black lines) and non-survivors (grey lines).

## DISCUSSION

In this study, we evaluated different iron-related parameters in acute COVID-19 positive and negative patients with similar age andprevalence of comorbidities. Iron- and immune-related determinations were made in the same samples, allowing the identification of correlations between some immune determinants and iron alterations. The first striking observation is the decrease in serum iron in COVID19-positive patients whencompared to COVID19-negative patients and healthy donors (Figure 1a) that has been reported in previous studies [6-8,10,13,17]. Here, we show that the reduction is clearly deeper than in patients with similar clinical manifestations but with a negative SARS-CoV-2 test. Interestingly, the levels of transferrin were also lower in COVID19-positive patients (Figure 1b), highlighting the negative acute phase nature of this protein. This parallel decrease in serum iron and transferrin may explain why transferrin saturation was not significantly lower in COVID19-positive patients than in COVID19-negative patients (Figure 1c). Decreased transferrin and transferrin saturation were found in COVID-19 patients in 3 previous studies [6,14,17], but not in another one [6]. Low circulating iron levels predicted a poor outcome. In fact, the patients with the lowest serum iron levels at first evaluation were the ones that died (Figure 2a). This result corroborates previous findings that associate lower serum iron levels with increased oxygenation demand, mild or severe respiratory failure [6,13] and fatal outcome [7]. Interestingly, we observed a tendency towards the stabilization of low serum iron over time in patients who died, while in those that recovered the serum iron levels tended to increase over time (Figure 5). This agrees with a previous study showing persistent alterations in iron status in severe cases of COVID-19 [13]. The low number of individuals observed precludes any definitive conclusion in this respect.

Despite the reduction of serum iron in COVID-19-positive patients in this cohort, we did not find important alterations in RBC, hemoglobin or hematocrit, in accordance with previous studies [6-8,13,21]. Increased heme was observed in sepsis patients [22] and in murine models of infection [23]. However, in our cohort, we did not find any increase in the levels of either heme or haptoglobin (Figure 1f,g), indicating that hemolysis is not common in COVID-19.

Globally, neutrophils, lymphocytes, monocytes, and platelets are lower in COVID-19-positive patients in our cohort. These data agree with other cohorts, except concerning neutrophils, which were described as increased [13].

In this study, the levels of circulating ferritin were clearly higher in COVID19-positive patients as compared, not only to healthy controls but also to COVID19-negative patients (Figure 1d). Other studies also found increased serum ferritin in patients with COVID-19 [7,13,20]. Some of those suggested the use of increased ferritin as a predictor of disease severity [11,13]. However, we could not find significant differences in the serum ferritin levels at admission between the different severity groups (Figure 2c). Interestingly, in the small number of patients followed over time, the ferritin levels at first evaluation tended to be higher but to decrease with time in those that recovered, as opposed to the fatal cases, who presented with slightly lower ferritin levels at diagnosis, but evolved to higher levels during hospitalization. These fluctuations in serum ferritin levels may explain variability in the values reported in different cohorts and are worth studying in more detail in future studies [6,13].

Decreased serum iron was not completely explained by increased hepcidin in our study, although it correlated with IL-6. The role of hepcidin in the reduction of circulating iron during SARS-CoV-2 infection is controversial [6,12,14]. Our data suggest that other cytokines, such as MCP-1, IL-8 and IL-18 may also modulating iron homeostasis, although the mechanisms are not explored. Importantly, along with low serum iron, high IL-6 levels were strongly associated with poor outcome. This confirms previous reports [24,25] and supports the recent World Health Organization (WHO) ecommendations for IL-6-receptor blockers as therapeutic strategy against COVID-19 [26,27].

This study has several limitations. Its retrospective and non-interventional nature limited the availability of follow-up samples with pre-established time-points. The lack of asymptomatic COVID19-positive patients precluded additional comparisons. Additionally, the COVID-19-negative individuals presented with heterogeneous clinical manifestations. Nevertheless, the large and well-defined cohort allowed the confirmation that COVID-19 causes more marked decreases in serum iron and increases in serum ferritin than other pathologies. More importantly, our data consolidate low serum iron, low transferrin, and high IL-6 as important predictors of disease severity in COVID-19.

## Data Availability

Raw data were generated at Centro Hospitalar Universitario Sao Joao and i3S - Instituto de Investigacao e Inovacao em Saude, Universidade do Porto. All data supporting the findings described in this study are available upon request from the corresponding author.

## Acknowledgements

The authors thank to all the staff at Clinical Pathology Department of CHUSJ, and the Tracy Core Facility of i3S, particularly Emília Cardoso, for technical assistance.

