## Supplementary data for "Iron related biomarkers predict disease severity in a cohort of Portuguese adult patients during COVID-19 acute infection"

Table S1– Distribution of COVID-19 patients among the severity groups, according to the presence of different comorbidities.

| Severity groups | 2 | 3 | 4 | 5 | Total |
|---|---|---|---|---|---|
| Count | 28 | 48 | 32 | 19 | 127 |
| Percentage in each group | 22.0 | 37.8 | 25.2 | 15.0 | 100 |
| **Diabetes** | | | | | |
| Count | 7 | 20 | 13 | 12 | 52 |
| Percentage in each group | 13.5 | 38.5 | 25 | 23.1 | 100 |
| Percentage within each severity group | 25.0 | 41.2 | 40.1 | 63.2 | |
| **Severity groups** | | | | | |
| $\chi^2$=6.883, p=0.077 | | | | | |
| **Hypertension** | | | | | |
| Count | 12 | 34 | 21 | 16 | 83 |
| Percentage in each group | 14.5 | 41.0 | 25.3 | 19.3 | 100 |
| Percentage within each severity group | 42.9 | 70.8 | 65.6 | 84.2 | |
| $\chi^2$=9.880, p=0.020 | | | | | |
| **Obesity** | | | | | |
| Count | 4 | 17 | 8 | 2 | 31 |
| Percentage in each group | 12.9 | 54.8 | 25.8 | 6.5 | 100 |
| Percentage within each severity group | 14.3 | 35.4 | 25.0 | 10.5 | |
| $\chi^2$=6.698, p=0.082 | | | | | |
| **Dyslipidaemia** | | | | | |
| Count | 7 | 21 | 15 | 13 | 56 |
| Percentage in each group | 12.5 | 37.5 | 26.8 | 23.2 | 100 |
| Percentage within each severity group | 25.0 | 43.4 | 46.9 | 13.0 | |
| $\chi^2$=8.805, p=0.032 | | | | | |
| **CKD** | | | | | |
| Count | 0 | 10 | 4 | 4 | 18 |
| Percentage in each group | 0 | 55.6 | 22.2 | 22.2 | 100 |
| Percentage within each severity group | 0 | 20.8 | 12.5 | 21.1 | |
| $\chi^2$=7.187, p=0.066 | | | | | |
| **CRD** | | | | | |
| Count | 5 | 8 | 2 | 4 | 18 |
| Percentage in each group | 27.8 | 44.4 | 11.1 | 16.7 | 100 |
| Percentage within each severity group | 17.9 | 16.7 | 6.3 | 21.1 | |
| $\chi^2$=2.225, p=0.522 | | | | | |
| **Anemia** | | | | | |
| Count | 0 | 4 | 0 | 0 | 4 |
| Percentage in each group | 0 | 100 | 0 | 0 | 100 |
| Percentage within each severity group | 0 | 8.3 | 0 | 0 | |
| $\chi^2$=6.797, p=0.079 | | | | | |
| **Hypocoagulation** | | | | | |
| Count | 1 | 8 | 2 | 0 | 11 |
| Percentage in each group | 9.1 | 72.7 | 18.2 | 0 | 100 |
| Percentage within each severity group | 3.6 | 16.7 | 6.3 | 0 | |
| $\chi^2$=6.842, p=0.077 | | | | | |

$\chi^2$: Pearson Chi-Square, p: Asymptotic significance (two-sided)

Table S2 – Counts and Percentage of first samples with detectable cytokines.*

| Cytokines (pg/ml) | COVID-19 Negative | COVID-19 Positive |
|---|---|---|
| IL-1beta | 14.5% (8/55) | 22.4% (12/49) |
| IL-6 | 74.5% (41/55) | 87.8% (43/49) |
| IL-8 | 96.4% (53/55) | 98.0% (48/49) |
| IL-10 | 78.1% (43/55) | 96.0% (47/49) |
| IL-12p70 | 60.0% (33/55) | 75.5% (31/49) |
| IL-18 [a] | 31.3% (15/48) | 59.5% (25/42) |
| IL-23[b] | 12.5% (6/48) | 33.3% (14/42) |
| IL-33 | 22.9% (11/48) | 23.8% (10/42) |
| MCP-1 | 100% (48/48) | 95.2% (40/42) |
| TNF alpha[c] | 12.7% (7/55) | 26.5% (13/49) |
| Interferon gamma[d] | 29.2% (19/48) | 76.1% (32/42) |
| Interferon alpha 2 | 54.1% (26/48) | 76.0% (32/42) |

*Inside brackets: number of positive samples/number of samples analysed.

COVID-19-positive vs COVID-19-negative [a]IL-18: 437.7±486.2 vs 912.2±806.7 pg/ml – p=0.0697;

[b]IL-23 10.89±13.07 vs 50.46±139.4 pg/ml – p=0.5466;

[c] TNF alpha 22.38±23.11 vs 30.92 ± 27.46 pg/ml – p=0.5880;

[d] Interferon gamma 17.28±43.28 vs 25.19±36.49 pg/ml – p=0.0062

Table S3 – Number of individual determinations per figure or table.

| | | COVID19 negative | COVID-19 positive | BD |
|---|---|---|---|---|
| | Iron | 84 | 114 | 35 |
| | Transferrin | 84 | 114 | 35 |
| | Tf sat | 84 | 114 | 35 |
| | Ferritin | 84 | 114 | 35 |
| | Hepcidin | 89 | 96 | 35 |
| | Heme | 142 | 85 | 35 |
| | Haptoglobin | 23 | 33 | 10 |
| | Erythropoetin | 65 | 99 | 35 |
| | RBC | 169 | 124 | |
| | Hb | 169 | 124 | |
| | HCT | 169 | 124 | |
| | MCV | 169 | 124 | |
| | MCH | 169 | 124 | |
| | MCHC | 169 | 124 | |
| | RDWCV | 169 | 124 | |
| | RDWSD | 169 | 124 | |
| | WBC | 169 | 124 | |
| | Neutrophils | 155 | 122 | |
| | Lymphocytes | 155 | 122 | |
| | Monocytes | 155 | 122 | |
| | platelets | 169 | 124 | |
| | platelets MPV | 166 | 121 | |
| | platelets PDW | 166 | 121 | |
| | Iron | | | |
| | severity 2 | 20 | 23 | |
| | severity 3 | 40 | 44 | |
| | severity 4 | 17 | 30 | |
| | severity 5 | 5 | 17 | |
| | Transferrin | | | |
| | severity 2 | 20 | 23 | |
| | severity 3 | 40 | 44 | |
| | severity 4 | 17 | 30 | |
| | severity 5 | 5 | 17 | |
| | Figure 1 and table 1 | | | |
| | Ferritin | | | |
| Figures 2 and 3 | severity 2 | 20 | 23 | |
| | severity 3 | 40 | 44 | |
| | severity 4 | 17 | 30 | |
| | severity 5 | 5 | 17 | |
| | WBC | | | |
| | severity 2 | 39 | 27 | |
| | severity 3 | 67 | 47 | |
| | severity 4 | 43 | 32 | |
| | severity 5 | 6 | 18 | |
| | Lymphocytes | | | |
| | severity 2 | 39 | 26 | |
| | severity 3 | 62 | 47 | |
| | severity 4 | 38 | 32 | |

**S1**

**a**

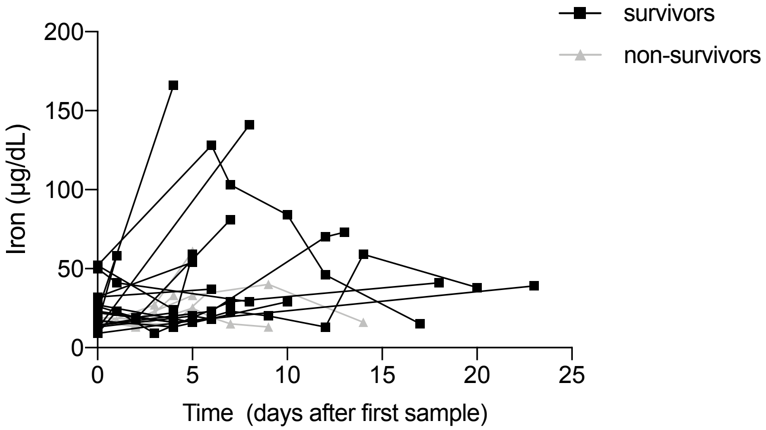
survivors
non-survivors
Iron (µg/dL)
0
50
100
150
200
0
5
10
15
20
25
Time (days after first sample)

**b**

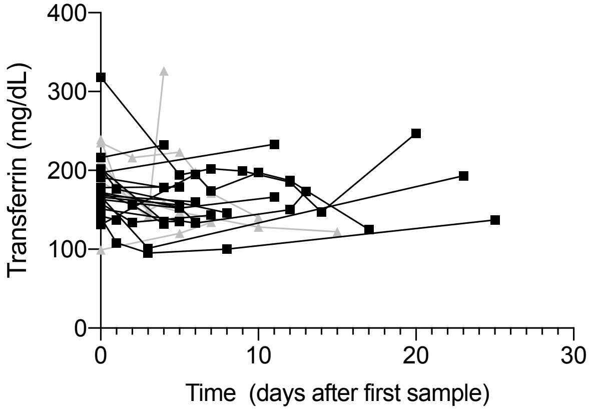
Transferrin (mg/dL)
0
100
200
300
400
0
10
20
30
Time (days after first sample)

**c**

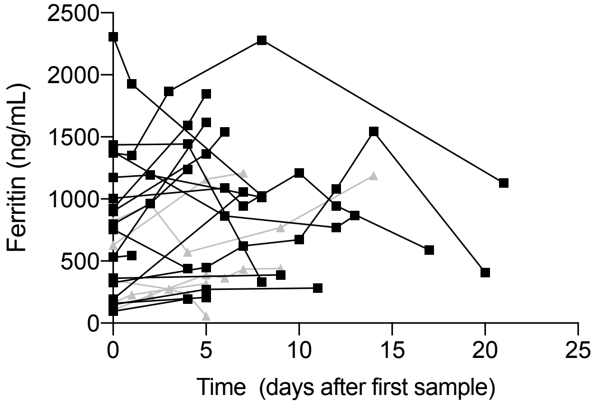
Ferritin (ng/mL)
0
500
1000
1500
2000
2500
0
5
10
15
20
25
Time (days after first sample)
